# Assessing Glucose Variability Metrics: Nerve Conduction Velocity in Children and Adolescents with Type 1 Diabetes

**DOI:** 10.1101/2025.03.25.25324553

**Authors:** Marc Robin Gruener, Erin West, Ute Muhitira, Katrin Heldt, Sarah Oberhauser, Dagmar l’Allemand, Mia Jovanova, Tobias Kowatsch, Philip Broser

## Abstract

**Background:** Diabetic peripheral neuropathy is a common microvascular complication of type 1 diabetes (T1D) that can manifest early in pediatric patients. While chronic hyperglycemia is an established risk factor, the role of glucose variability in the development of neuropathy remains controversial, particularly in children and adolescents.

**Methods:** In this single-center retrospective cross-sectional study, we evaluated associations between height-adjusted nerve conduction velocity (aNCV) and multiple glucose variability metrics in children and adolescents with type 1 diabetes. We analyzed continuous glucose monitoring data from the 90 days preceding standardized neurophysiological assessment to calculate established glucose variability metrics (SD, CONGA, ADRR, MAG, GVP) alongside our novel Glucose Fluctuation Moment Index (GFMI). The GFMI uniquely integrates both the velocity of glucose changes and their distance from the glycemic target. Using linear regression models adjusted for diabetes duration, we assessed the relationship between each glucose variability metric and aNCV, employing leave-one-out cross-validation to evaluate predictive accuracy despite limited sample size.

**Results:** Among 42 eligible participants (mean age 12.2 ± 3.2 years, diabetes duration 4.1 ± 3.2 years), mean aNCV was negative in the peroneal nerve (-2.9 ± 2.9 m/s). Linear regression models adjusting for diabetes duration revealed no statistically significant associations between any glucose variability metrics (SD, CONGA1, CONGA24, ADRR, MAG, GVP, or GFMI) and peroneal nerve aNCV (all p>0.05). In predictive modeling using leave-one-out cross-validation, the Glycemic Variability Percentage (GVP) demonstrated the best predictive performance (RMSE 2.99 m/s with all observations, 2.46 m/s after removing three identified outliers), but offered minimal improvements (4.28 % and 6.08 %) over the baseline- (simply predicting the mean aNCV) or the glucose SD-based model. Outlier analysis revealed notable clinical factors potentially affecting nerve function, including physical activity.

**Conclusions:** While early neurophysiological changes were observed in our pediatric T1D cohort, neither established glucose variability metrics nor our novel GFMI demonstrated superior predictive accuracy for the peroneal aNCV compared to the glucose SD, the clinically established GV measure, nor achieved clinically relevant accuracy. Further research with larger cohorts and longitudinal designs is needed to account for relevant confounding variables like C-peptide and physical activity to better understand these complex relationships.

## 1 Introduction

### 1.1 Clinical Significance and Impact of Diabetic Peripheral Neuropathy

Diabetic peripheral neuropathy (DPN) is the most common microvascular complication of diabetes, affecting nearly half of diabetic patients over their lifetime.(1–4) DPN presents as peripheral nerve dysfunction and is often characterized by a “stocking and glove distribution”, primarily affecting the lower limbs and hands. Symptoms include poor balance, pain, and reduced sensory perception.(5)

In pediatric populations with type 1 diabetes (T1D), DPN prevalence estimates range from 7% to 57% or higher, depending on diagnostic criteria and patient characteristics.(6) Longitudinal studies highlight its early onset and progressive nature. For instance, Hajas et al. (7) observed an increase in DPN from 24.2% to 62.9% over 10 years in young individuals with T1D, while Hyllienmark et al. (8) reported a 15% incidence over five years. These findings underscore the need to identify modifiable risk factors and predictive markers to intervene before irreversible nerve damage occurs.

Early detection of neurophysiological changes and identification of modifiable risk factors are essential for implementing preventive interventions, before irreversible nerve damage occurs.

### 1.2 Pathophysiological Mechanisms of Nerve Damage in Diabetes

Saltatory action potential conduction in peripheral nerves depends critically on stable metabolic homeostasis. Diabetes disrupts this stability through multiple pathways. Chronic hyperglycemia activates damaging biochemical processes, including polyol pathway activation, advanced glycation end-product formation, oxidative stress, and inflammation, leading to axonal degeneration and demyelination.(5)

Beyond chronic hyperglycemia, emerging evidence suggests that glucose fluctuations may independently contribute to neuronal damage. Glycemic variability (GV) induces oxidative stress more potently than sustained hyperglycemia.(9) Rapid shifts between hyperglycemia and normoglycemia, something most individuals affected by T1D experience, trigger cellular mechanisms that increase reactive oxygen species production and upregulate inflammatory pathways.(9–11) This instability may expose peripheral nerves to recurrent metabolic stress, contributing to functional neuropathic damage.

### 1.3 Relationship Between Glycemic Variability and Diabetic Neuropathy: Conflicting Evidence

The relationship between glycemic variability and diabetic neuropathy remains controversial. Previous research by the team found that GV, measured by glucose standard deviation, exhibits statistically significant negative correlations with NCV in children and young adults with T1D.(12,13) Similar findings have been reported by others, who demonstrated associations between various measures of GV and neuropathic outcomes.(14,15)

However, a substantial body of evidence suggests no significant relationship between GV and neuropathic complications. Evidence linking variability to neuropathy is inconsistent.(16) Particularly relevant to our present work, Christensen et al. (17) found that initial associations between variability metrics and nerve conduction parameters in young adults with T1D disappeared after adjustment for confounders.

### 1.4 Limitations of Current Glucose Variability Metrics and Proposal of the Glucose Fluctuation Moment Index (GFMI)

Numerous metrics have been proposed to quantify GV, including standard deviation (SD), coefficient of variation (CV), mean amplitude of glycemic excursions (MAGE), continuous overall net glycemic action (CONGA), average daily risk range (ADRR), mean absolute glucose change (MAG), and glycemic variability percentage (GVP). Each has limitations in capturing clinically relevant glycemic patterns.(18,19)

SD does not differentiate between the order of glucose values.(20) GVP compares the length of the glucose trace to a horizontal line but ignores the overall glucose levels. Other metrics emphasize different mathematical properties of GV but lack a comprehensive evaluation of their systematic associations with biological markers of tissue damage in studies and relevant clinical endpoints.

To address these limitations, we propose the Glucose Fluctuation Moment Index (GFMI), which combines the velocity of glucose changes with the distance from the target range. The theoretical foundation and calculation method of the GFMI will be presented in the Methods section.

### 1.5 Study Objectives and Hypotheses

This study aims to:

1. Establish reference values for various GV metrics in pediatric individuals with T1D.
2. Assess the relationship between GV metrics and aNCV as a clinical endpoint for DPN in pediatric individuals with T1D.
3. Evaluate the predictive accuracy of GV metrics for the estimation of aNCV.

We hypothesize that metrics capturing rapid changes at extreme values (particularly GFMI) will show negative linear relationships with aNCV and higher predictive accuracy for aNCV compared to the glucose SD. Our findings may help refine risk assessment and early intervention strategies for DPN in pediatric individuals with T1D.

## 2 Methods

### 2.1 Study design

This single-center retrospective cross-sectional study was conducted at the Children’s Hospital of Eastern Switzerland (OKS), representing a collaboration between the endocrinology/diabetology and neurophysiology departments. The local ethics committee approved the study (St. Gallen, Switzerland; EKOS 22/018) and the study is registered with the Swiss project database (2022–00216). Prior to inclusion, written informed consent was obtained from legal guardians and patients (for those ≥14 years).

### 2.2 Participants & Recruitment

Children and adolescents with T1D were consecutively enrolled between June 2019 and February 2025. T1D diagnosis was confirmed through autoantibody measurement (GAD, ZnT8, IA2, ICA, and/or IA), clinical presentation, C-peptide levels, and genetic analyses where indicated. We excluded patients <5 years of age as their nerve conduction measurements would not be comparable due to significant developmental changes in the nervous system during early childhood.(21,22) Additionally, patients with other non-diabetes related chronic diseases, premature birth, or a family history of inherited neurological diseases were excluded. Patients not using continuous glucose monitoring (CGM) devices as part of their diabetes therapy or with less than 70% of CGM values available were also excluded per established protocols.(23)

All eligible patients underwent standardized neurophysiological assessment at the OKS neurophysiology unit (22), with only the initial nerve conduction study (NCS) considered for this cross-sectional analysis. For patients with multiple NCS observations meeting these criteria, we included only the first observation to maintain statistical independence and prevent the influence of a potential bias induced by knowledge about previous measurements or changes to the therapy in response to a previous NCV measurement.

### 2.3 Variables + Data Sources

#### 2.3.1 Nerve Conduction Study

Nerve conduction studies were performed by trained medical professionals following a standardized protocol.(22) The protocol measured NCV in four standardized nerves: three motor nerves (peroneal, tibial, and median) and one sensory nerve (median). All measurements were conducted in a temperature-controlled environment on the left extremities (right side used only when left-side measurement was not feasible).

To account for the known negative association between NCV and body height (24), we calculated height-adjusted NCV (aNCV) values using a linear regression model in accordance with Hyllienmark et al.(24), using data from 65 healthy subjects at the OKS.(12)

#### 2.3.2 Laboratory Data

Blood samples for HbA1c was collected on the same day as the NCS and analyzed using the Abbott AFINION™ 2. While these laboratory measurements provide important point-in-time assessments of glycemic control, CGM offers detailed insights into daily glucose dynamics.

#### 2.3.3 Continuous Glucose Monitoring Data

CGM data were obtained from participants using various systems (FreeStyle Libre 2/3, Dexcom G6/7, and Medtronic Guardian 3/4) accessed through manufacturer-specific platforms (Medtronic CareLink, Dexcom Clarity, Abbott LibreView) or Glooko. We analyzed a 90-day window preceding each participant’s nerve conduction study based on established evidence that this timeframe: 1) aligns with the clinical interpretation of HbA1c representing ∼3 months of glycemic control (23), 2) provides sufficient data for reliable calculation of variability metrics (25), and 3) represents a clinically relevant period during which glycemic patterns could influence nerve function.

Our processing protocol standardized glucose measurement ranges (2.2-22.2 mmol/L) with values outside this range coded as missing to address manufacturer-specific differences. All measurements in mg/dL were converted to mmol/L. We resampled all CGM time series to standardized 15-minute intervals using forward-filling (max 1 period) to account for varying measurement frequencies across sensor models. Periods of sensor changes or warm-ups were retained as missing values.

All CGM readings were collected in their respective time zones as recorded by the manufacturer platforms, with timestamps verified for consistency during processing. No missing values were imputed to maintain data integrity and reflect real-world CGM use patterns.

#### 2.3.4 Glucose Variability Metrics

Previous research has demonstrated that standard glycemic control measures may not fully capture the dynamic nature of glucose fluctuations that impact clinical outcomes.(10,26) While SD of glucose has shown associations with reduced nerve conduction velocity (12), it cannot distinguish between clinically distinct glucose patterns, particularly regarding the timing and magnitude of changes relative to target glucose levels (20).

Based on a comprehensive literature review by Donaldson et al. (19), we selected four established glucose variability metrics that capture distinct aspects of glycemic control. Each metric was chosen for its validated relationship with clinical outcomes and its unique approach to quantifying glucose dynamics:

The standard deviation (SD) of glucose measurements represents the most fundamental measure of glycemic variability, quantifying the average deviation from the mean glucose level. While straightforward to calculate and widely used in clinical practice (27), SD treats all deviations equally, regardless of their order, clinical significance, or their relationship to target glucose levels.

The Continuous Overall Net Glycemic Action (CONGAn) evaluates intraday glycemic variation by analyzing differences between glucose readings at specified time intervals. This metric is particularly valuable for assessing temporal glucose dynamics and has been validated in pediatric Type 1 diabetes populations. CONGA offers flexibility in time interval selection, allowing analysis of both short-term and long-term glucose patterns.(18) For our analysis, we focus on CONGA1 and CONGA24 to reflect short-term fluctuations and day-to- day patterns.

The Average Daily Risk Range (ADRR) addresses the asymmetric nature of glucose variations by incorporating separate risk functions for hypo- and hyperglycemia. Through logarithmic transformation of glucose values, ADRR provides a balanced assessment of extreme glucose events, making it particularly relevant for evaluating overall glycemic risk.(28–30)

The Mean Absolute Glucose Change (MAG) quantifies the cumulative magnitude of glucose changes over time. This metric effectively differentiates between glucose profiles that may have similar means or standard deviations but represent distinctly different patterns of glycemic control. MAG has demonstrated a particular utility in identifying clinically significant glucose variations in intensive care settings.(20)

The Glycemic Variability Percentage (GVP) takes a geometric approach by comparing actual glucose trace lengths to minimal possible lengths. This method specifically penalizes high-frequency and high-amplitude oscillations, providing insight into glucose stability that complements traditional variability measures.(31)

To address limitations in existing metrics, we propose the Glucose Fluctuation Moment Index. This metric combines two key components: the squared velocity of glucose concentration changes (the discrete first derivative of the glucose timeseries) weighted by the squared distance from the glycemic target of 5 mmol/L:

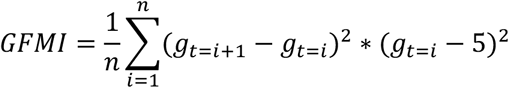

The GFMI was designed to emphasize large glucose changes occurring far from the target range, based on our hypothesis that such excursions may have particularly detrimental effects on nerve vitality. A comprehensive overview of the different metrics can be found in Table 1, including the purpose, mathematical formulas, selected examples of clinical and scientific application, and distinguishing features.

**Table 1:**
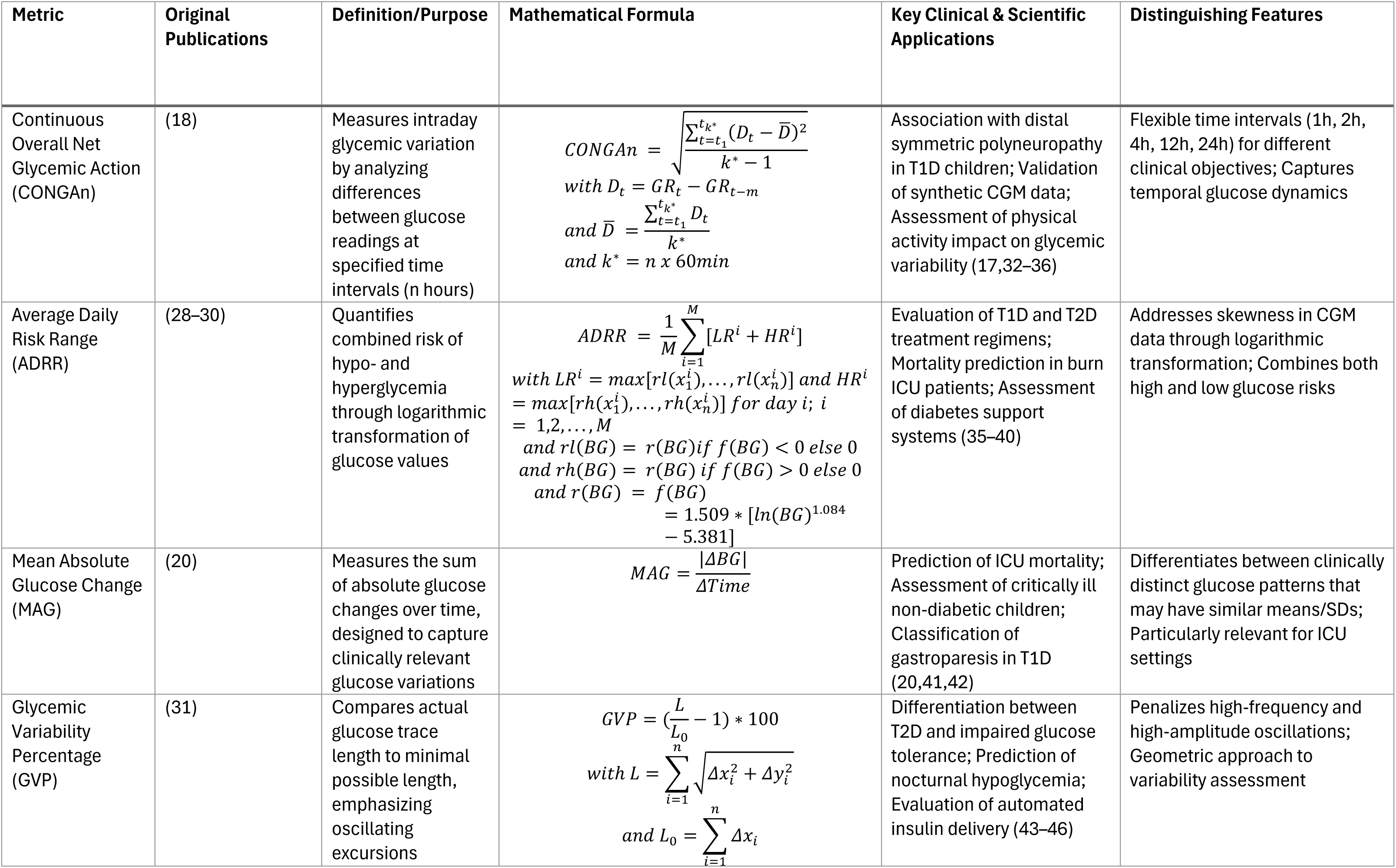
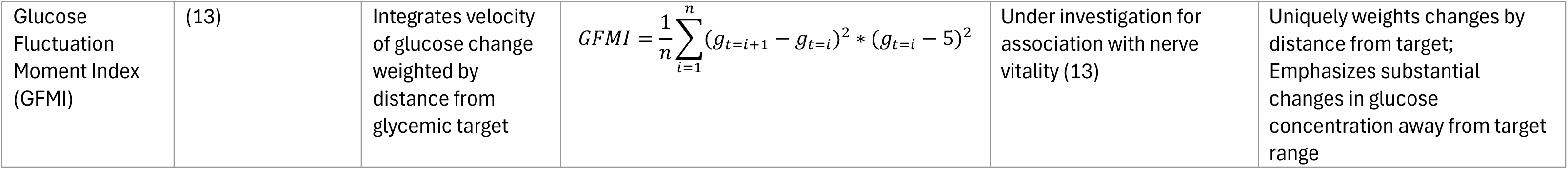
Overview of selected Glucose Fluctuation Metrics.

Following data preprocessing as described earlier, we calculated this comprehensive set of digital biomarkers (SD, GFMI, CONGAn, ADRR, MAG, and GVP) from the CGM data to provide a thorough assessment of glycemic control, with particular emphasis on glucose concentration fluctuations and variability. Each metric offers insights into distinct aspects of glycemic control, allowing for a more nuanced understanding of glucose dynamics than any single measure alone.

#### 2.3.5 Statistical Methods

To evaluate relationships between GV metrics and aNCV, we employed two complementary analytical approaches. First, we assessed statistical relationships between each GV metric and aNCV of the peroneal nerve, chosen based on previous research (12), while adjusting for diabetes duration (16,17), using linear regression models. We calculated GV coefficient estimates, 95% confidence intervals, and p-values for each model, testing the null hypothesis *β_GV Metric_* = 0. We identified potential statistical outliers using studentized residuals with a threshold at the two-sided 95% t-distribution quantile. Each identified outlier was subsequently reviewed by a certified endocrinologist and a neurophysiologist to analyze the cases from both medical disciplines and determine whether there were clinical explanations for the statistical deviation. To assess the robustness of our findings, we report all subsequent analyses pre- and post-removal of our identified outliers.

Second, we calculate the predictive accuracy, leveraging linear regression models combining diabetes duration with individual glucose variability metrics. Given our limited sample size, we employed leave-one-out cross-validation (LOO-CV) to maximize the training data while still obtaining reliable estimates for out-of-sample prediction accuracy. We evaluated model performance using root mean squared error (RMSE).

All models followed a consistent structure, incorporating one glucose variability metric (CONGA1, CONGA24, ADRR, MAG, GVP, or GFMI) and diabetes duration as predictors of an NCV:

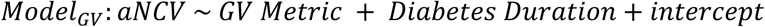

We also included two comparison models: a baseline model using only the mean aNCV, and a standard-of-care (SOC) model combining glucose SD with diabetes duration:

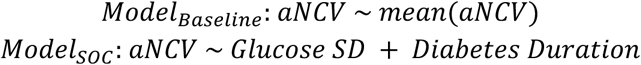

To ensure model validity, we conducted comprehensive diagnostic analyses, including examination of predicted versus actual values and residual plots for each regression model.

We implemented our statistical analyses using two programming environments: R v4.3.2 with the lme4 package v1.1-35.3 for association analyses, and Python 3.10 with pandas v2.0.3, scipy v1.11.4, statsmodels v0.14.0, and sklearn v1.3.0 for predictive modeling.

## 3 Results

### 3.1 Patient Selection and Demographic

We identified 42 eligible patients with sufficient data for analysis from 138 NCS performed in our center’s T1D population (Figure 1). Exclusions were primarily due to insufficient CGM data (<70% over 90 days; n=71), age <5 years (n=13), or repeated measurements (n=9).

**Figure 1:**
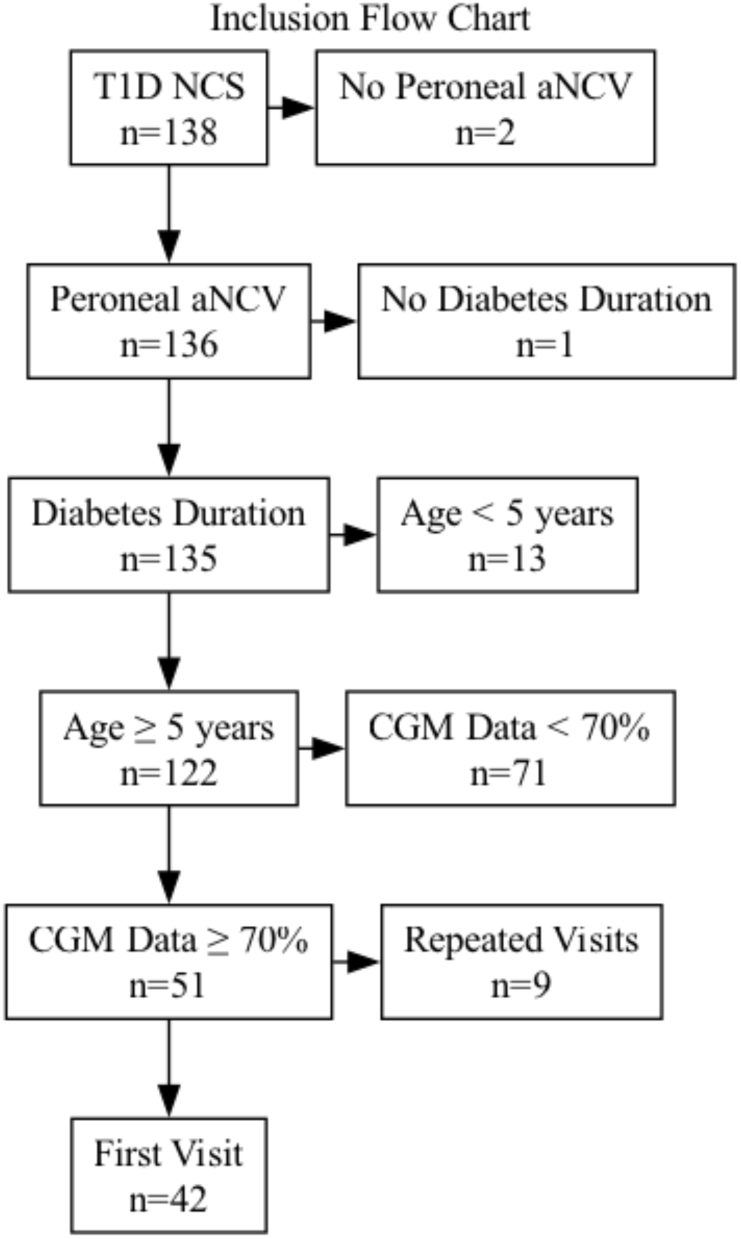
Inclusion Flow Chart.

The study sample (Table 2) had a mean age of 12.2 ± 3.2 years [mean ± SD] and 47.6% female participants. Mean height and weight percentiles were 70.1 ± 21.8% and 66.9 ± 23.8% respectively. Mean diabetes duration was 4.1 ± 3.2 years with a mean HbA1c of 7.4 ± 1.2%, indicating suboptimal control according to the current pediatric diabetes guidelines of 6.5 or 7.0% (47).

**Table 2:** Characteristics of Participants.

| Demographic Data | Value (n=42) |
| --- | --- |
| Age, years, mean ± SD | $12.2 \pm 3.2$ |
| Female, n (%) | 20 (47.6%) |
| Height, cm, mean ± SD | $155.1 \pm 19.5$ |
| Height percentile, %, mean ± SD | $70.1 \pm 21.8$ |
| Weight, kg, mean ± SD | $49.9 \pm 18.3$ |
| Weight percentile, %, mean ± SD | $66.9 \pm 23.8$ |

| **Diabetes Characteristics** |  |
| --- | --- |
| Age at diagnosis, years, mean ± SD | 8.2 ± 3.7 |
| Diabetes duration, years, mean ± SD | 4.1 ± 3.2 |
| HbA1c, %, mean ± SD | 7.4 ± 1.2 |

Participants used the following CGM devices: Dexcom G6/G7 (n=13, 31.0 %), Medtronic Guardian 3/4 (n=13, 31.0 %), and Abbott FreeStyle Libre 2/3 (n=16, 38.1 %).

### 3.2 Primary Measures

#### 3.2.1 Diabetes-related Measures

In our final analysis data set, mean CGM data availability was 90.5 ± 8.8% across the 90- day pre-NCS observation window. Standard glycemic metrics revealed average glucose levels of 9.1 ± 1.5 mmol/L, and suboptimal time-in-range (62.8 ± 14.9 % within 3.9-10.0 mmol/L) compared to the recommended clinical target of >70% (48). Mean glucose standard deviation was 3.6 ± 0.7 mmol/L.

In Table 3, MAG value (2.88 ± 0.47 mmol/L/h) reflected the average hourly glucose change magnitude, while GVP (23.65 ± 7.57 %) quantified the geometric variability of glucose traces. Mean ADRR was 42.32 ± 9.42, quantifying risk from combined hypo- and hyperglycemic excursions. The GFMI metric demonstrated considerable between-patient variation (43.05 ± 27.68).

**Table 3:**
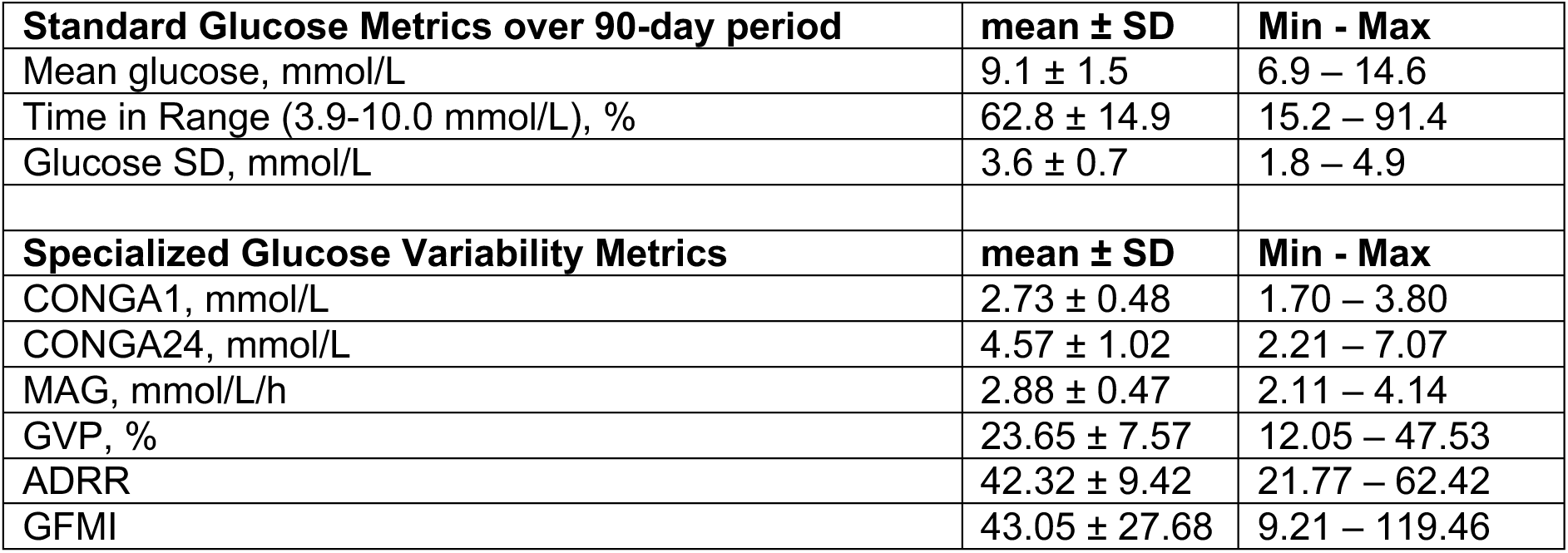
Glucose Fluctuation Indices.

#### 3.2.2 Peripheral Nervous System-related Measures

Using the formula for the aNCV based on the healthy population, we find, on average, negative aNCV in all four nerves for the diabetic sample (Table 4). The lowest mean aNCV was observed in the median sensory nerve (-3.2 ± 5.1 m/s), followed by the peroneal (-2.9 ± 2.9 m/s), tibial (-3.0 ± 4.6 m/s), and median motor nerves (-1.5 ± 3.7 m/s).

**Table 4:** Nerve Conduction Velocity Results.

| Nerve | n | NCV<br>m/s, mean ± SD | aNCV<br>m/s, mean ± SD |
| --- | --- | --- | --- |
| Peroneal | 42 | 49.2 ± 3.8 | -2.9 ± 2.9 |
| Tibial | 41 | 48.1 ± 4.6 | -3.0 ± 4.6 |
| Median (motor) | 42 | 56.8 ± 3.6 | -1.5 ± 3.7 |
| Median (sensory) | 42 | 53.7 ± 5.3 | -3.2 ± 5.1 |

### 3.3 Main Results

#### 3.3.1 Relationships between GV Metrics and aNCV

Figure 2 illustrates the relationships between six key GV metrics and aNCV, with points colored by diabetes duration (<5 years in grey, ≥5 years in black).

**Figure 2:**
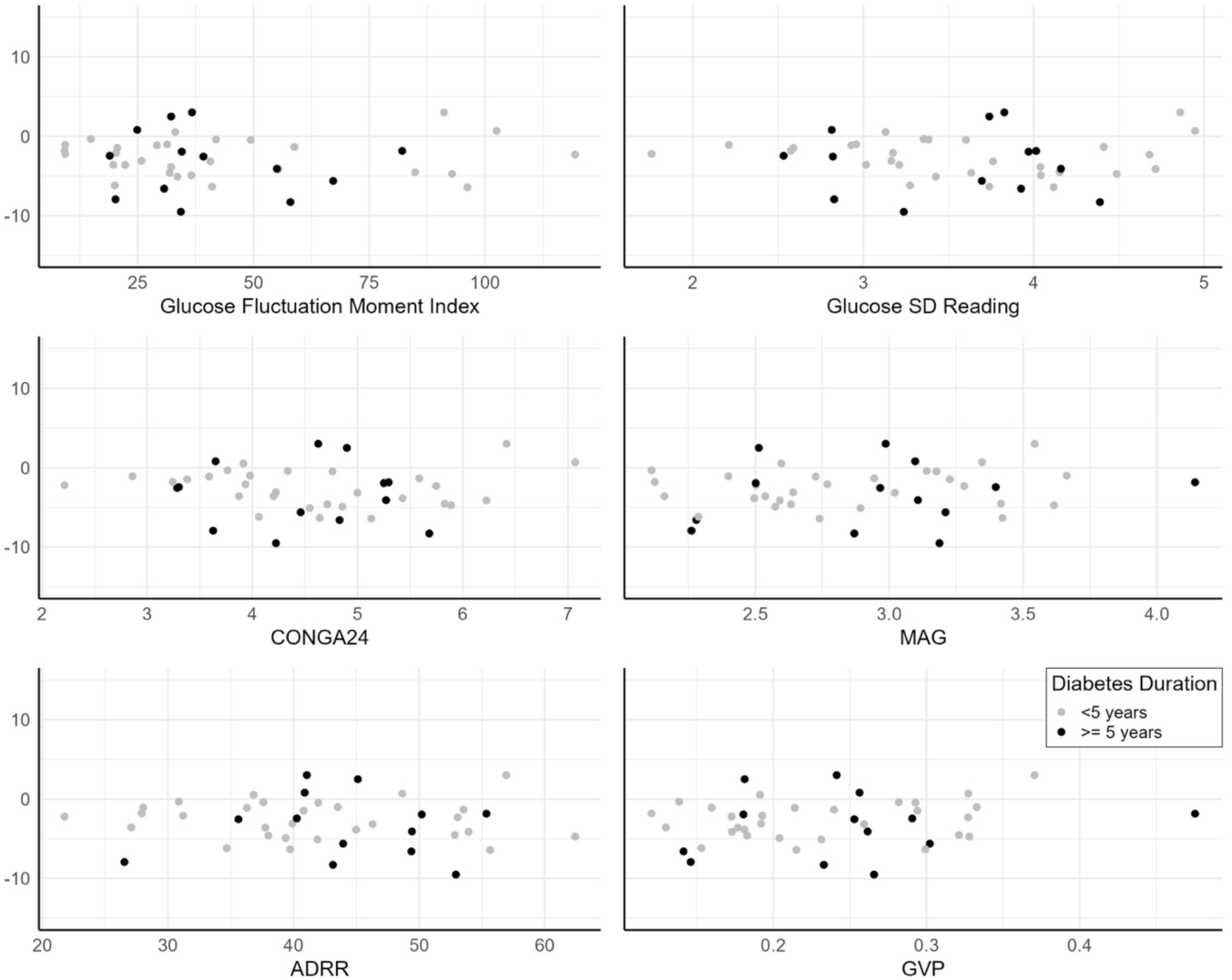
Scatterplots of the Adjusted Peroneus Nerve Conduction Velocity against selected Glucose Fluctuation Metrics.

In none of the GV-based models, adjusted for diabetes duration, did the GV coefficients statistically significantly differ from 0 (all p > 0.05, Table 5).

**Table 5:** Glucose-Fluctuation based Linear Models of the Peroneus aNCV.

| Model | Coefficient | 95% CI | p-value |
| --- | --- | --- | --- |
| Glucose SD (SOC) | -0.147 | -1.413, 1.19 | 0.816 |
| CONGA1 | 0.980 | -0.952, 2.912 | 0.311 |
| CONGA24 | -0.049 | -0.961, 0.862 | 0.914 |
| GFMI | 0.000 | -0.034, 0.034 | 0.994 |
| ADRR | -0.049 | -0.113, 0.087 | 0.800 |
| MAG | 1.018 | -0.948, 2.985 | 0.301 |
| GVP | 7.850 | -4.320, 20.019 | 0.200 |

Normality of residuals was confirmed by visual analysis of Q-Q-plots.

#### 3.3.2 Identification of Outliers

Our analysis identified three patients (7.1 % of the sample) as statistical outliers across all regression models, with studentized residuals exceeding the two-sided 95% t-distribution threshold. Two of the patients (ID 17 & 22) displayed aNCV for the peroneal nerve above the expected value for a healthy individual of their height (heights: 1.35 & 1.80 m) with a NCV of 57 and 52 m/s and an aNCV of 3.02 and 2.50 m/s respectively. The NCV and aNCV for the third patient (ID 33, height: 1.30 m) were recorded at 45 and -9.51 m/s. Brief case reports for all three individuals are provided in section 4.4. The following evaluation of the predictive model accuracy was conducted before and after removal of the three identified outliers.

#### 3.3.3 Predictive Accuracy

To evaluate the clinical utility of these metrics beyond statistical relationships, we assessed their predictive performance using leave-one-out cross-validation, reporting the RMSE in Figure 3. The baseline model yielded an RMSE of 2.97 m/s with all observations included and 2.57 m/s after removal of outliers. Among the glucose variability metrics, GVP demonstrated the best predictive accuracy (RMSE 2.99 m/s with all observations, 2.46 m/s without outliers), representing almost equal performance with the baseline model across all observations and only a modest 4.28 % improvement when comparing the results after outliers were removed.

**Figure 3:**
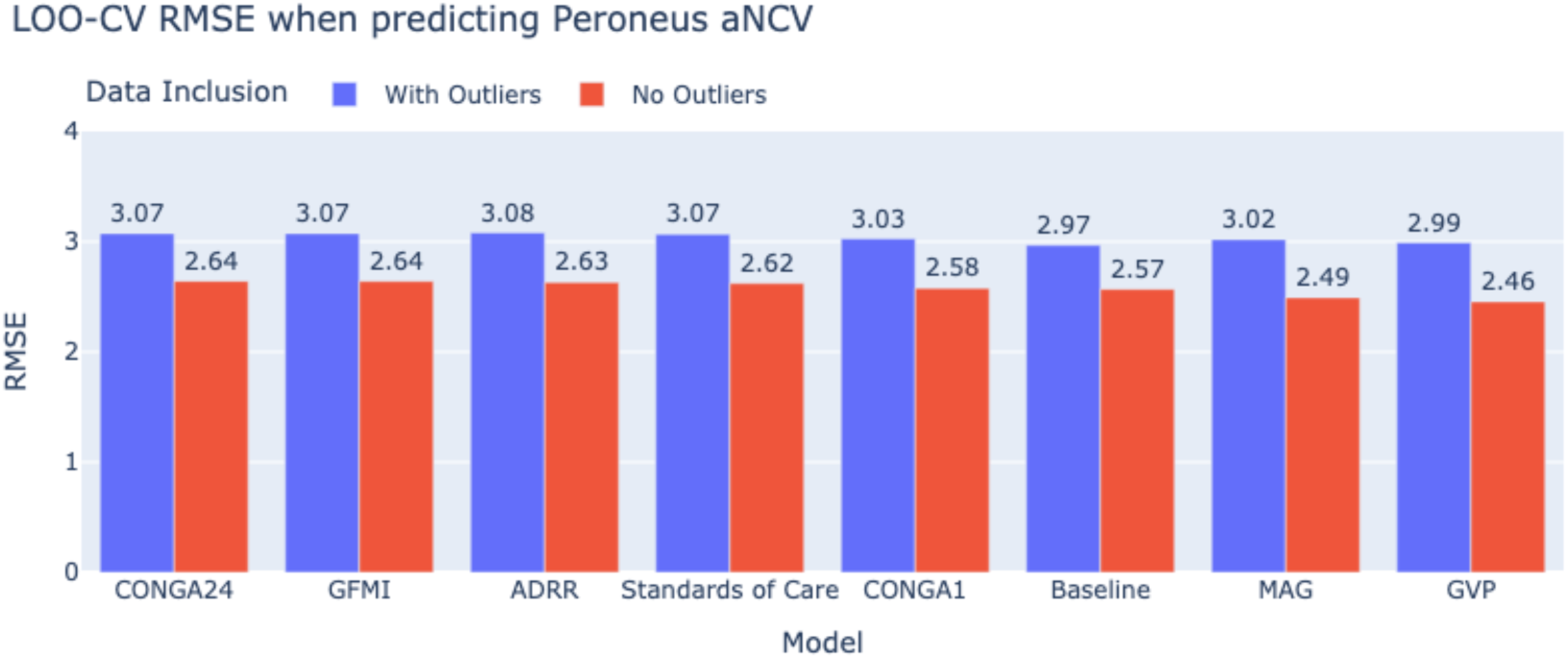
Average LOO-Cross Validation RMSE when predicting adj Peroneus NCV based on different Glucose Fluctuation Metrics and the Baseline Model.

After removing outliers, RMSE decreased by approximately 15.1 % across all models. The accuracy ranking of the different GV metrics remained almost consistent, with GVP and MAG showing the most favorable predictive accuracy. The only difference in the ranking was the Baseline model, scoring first place with all data points and third after removal of outliers, and the ADRR model moving from the last to the sixth place.

### 3.4 Analysis of Outliers

Three patients of the 42 patients were classified as statistical outliers. A closer examination of their medical records revealed the following findings:

Patient 17 (male; height: 1.35 m; NCV: 57 m/s; aNCV: 3.02 m/s) regularly plays field hockey and is also highly active in other sports. Besides diabetes, he has no other known medical conditions.

Patient 22 (male; height: 1.80 m; NCV: 52 m/s; aNCV: 2.50 m/s) suffers from coeliac disease and rheumatoid arthritis. He was training for a marathon and was also intensively active in additional sports.

Patient 33 (male; height: 1.30 m; NCV 45 m/s; aNCV: -9.51 m/s) one-sided nephropathy but otherwise normal kidney function. T1D was diagnosed abroad before moving to Switzerland. Previous indication for DPN.

## 4 Discussion

### 4.1 Summary of Key Findings and Contributions

Our cross-sectional study provides novel insights into the relationship between GV metrics and early neurophysiological changes in children and adolescents with T1D. We found consistently negative mean aNCV across all measured nerves. Despite comprehensive analysis of multiple GV metrics—including the Glucose Fluctuation Moment Index (GFMI)— none demonstrated a statistically significant relationship with aNCV after adjusting for diabetes duration, nor did they achieve clinically meaningful predictive accuracy.

This work contributes to the field in three ways. First, we move beyond simple correlation analysis to evaluate the actual predictive performance of GV metrics against a clinically relevant neurophysiological outcome, establishing a higher standard for assessing their utility. Second, we provide detailed nerve conduction data in a pediatric population with T1D where normative values are scarce, offering valuable reference information for clinical interpretation. Third, we introduce the GFMI, which captures glycemic excursions weighted by their distance from target, conceptually addressing limitations of existing metrics.

Our predictive modeling results demonstrate that none of the GV metrics achieved clinically actionable prediction accuracy. With RMSE values nearly identical across models (RMSE 2.97 to 3.08 m/s and 2.46 to 2.64 m/s after outlier removal), all models fell short of the minimal clinically important difference for NCV of approximately 1.2 m/s, however, this number is debated (49) and might not be applicable for children and adolescents. These findings do not support adopting complex GV metrics or the glucose SD and diabetes duration alone as predictors for peroneal aNCV in clinical practice. Additionally, we found no reason to prioritize advanced GV metrics like ADRR and GFMI over the established glucose SD, given their additional technical complexity, lack of established reference ranges, especially for children and adolescents, and educational burden for clinicians.

### 4.2 Comparison with Previous Research

#### 4.2.1 Neurophysiological Findings

The negative mean aNCV observed in our cohort aligns with existing evidence that first signs of diabetic neuropathy can be found in pediatric populations.(7,8,50,51)

Our height-adjusted approach to NCV interpretation represents a methodological advancement over studies using unadjusted values. By accounting for the known relationship between height and NCV, we address a critical confounder in pediatric populations experiencing rapid growth. This approach enhances the accuracy of detecting diabetes-related neurophysiological changes independent of developmental factors.

#### 4.2.2 Glucose Variability and Neuropathy: Contradictory Evidence

The relationship between GV and diabetic peripheral neuropathy remains uncertain, with conflicting findings throughout the literature. Several studies, including previous work by our team (12,13), have found significant associations between glucose fluctuations and neuropathic outcomes. For instance, Xu et al. (14) reported that increased GV measured by CGM correlated with peripheral nerve dysfunction independent of HbA1c in adults with type 2 diabetes. Similarly, Akaza et al. (15) found that glucose variability (mean amplitude of glycemic excursions; MAGE) “was independently associated with a higher risk of medial plantar neuropathy” in individuals affected by T1D and T2D.

However, an equally compelling body of evidence indicates no significant relationship between GV and neuropathic complications. Siegelaar et al. (16) concluded that while GV is not an independent risk factor for DPN after adjusting for HbA1C or mean glucose based on their analysis of the Diabetes Control and Complications trial.

Our findings align closely with the cross-sectional study by Christensen et al. (17), which examined the same relationship in young adults (18-24 years) with T1D. Their investigation, involving 133 participants and a comprehensive assessment of peripheral and autonomic neuropathy, mirrors our methodological approach while targeting a slightly older but developmentally adjacent population. Despite their larger sample size, Christensen and colleagues found that initial associations between variability metrics (coefficient of variation, SD, MAGE, CONGA) and nerve conduction parameters disappeared after adjustment for potential confounders and multiple testing. Their conclusion that “GV may not be a risk factor for early diabetic neuropathy in young adults with type 1 diabetes” remarkably parallels our findings in pediatric patients, suggesting consistency across developmental stages from childhood through early adulthood.

A fundamental challenge in interpreting this contradictory literature lies in methodological inconsistencies across studies. Diagnostic criteria for diabetic peripheral neuropathy vary substantially, with some research relying on symptom questionnaires, others on quantitative sensory testing, and others—like our study—on nerve conduction assessments. Each approach captures distinct aspects of nerve dysfunction, from subjective symptoms to small or large fiber function. Similarly, the definition and quantification of GV varies considerably between studies. Researchers employ diverse metrics ranging from simple standard deviation to complex algorithms like CONGA, MAGE, and our novel GFMI, each highlighting different mathematical properties of glycemic fluctuations. Without standardized approaches to both neuropathy assessment and glucose variability calculation, direct comparison across studies remains problematic, potentially explaining some of the contradictory findings in the literature.

Our study addresses these challenges through rigorous methodology, including height-adjusted nerve conduction measurements, the gold standard, and a comprehensive assessment of multiple glucose variability metrics using a standardized data processing protocol. Nevertheless, our findings join others in questioning whether current GV metrics effectively capture the aspects of glycemic control most relevant to neurological health in pediatric diabetes patients.

### 4.3 Strengths and Limitations

#### 4.3.1 Strengths

Our study features several methodological strengths that enhance its contribution to the field. By focusing on a pediatric population with T1D, we address an understudied population where early intervention could potentially prevent irreversible nerve damage. Our analytical approach using leave-one-out cross-validation to assess predictive accuracy represents a significant methodological improvement over simple correlation analyses that dominate the literature. Additionally, our rigorous height-adjustment methodology for NCV interpretation addresses a crucial confounder in growing children, while our standardized CGM data processing protocol ensures comparability across different sensor systems—an important consideration as CGM technology evolves.

#### 4.3.2 Limitations

Several important limitations must be considered when interpreting our findings. After applying stringent exclusion criteria, our relatively small sample size (n=42) limits our ability to draw conclusive findings. Furthermore, our study population represents only patients who consistently use CGM devices and met our inclusion criteria (only 42 of 138 eligible patients), potentially introducing selection bias.

No power calculation was performed prior to conducting the study due to the lack of comparable research focusing on the peroneal aNCV and advanced GV metrics in a pediatric population. Therefore, our findings should be interpreted with caution.

Additionally, the technical limitations of CGM technology constrain our analysis. Even the most modern systems have a measurable error of 9.7 – 16.6 % MARD (52), introducing noise that may obscure subtle relationships with neurophysiological outcomes. When glucose values exceed measurement ranges, researchers face a methodological dilemma: exclude these out-of-range periods or impute values. This mainly affects patients with the poorest control—precisely those who might be at the highest risk for neuropathy—potentially underrepresenting relationships between extreme GV and nerve damage.

### 4.4 Future Directions

Our findings highlight three priority areas for future research. First, studies with larger samples are needed to establish whether specific GV metrics are associated with early neurophysiological changes in pediatric populations with T1D.

Second, longitudinal cohort studies with repeated neurophysiological assessments would provide crucial insights into the temporal relationship between glycemic patterns and nerve function. Such studies could identify early warning signs and establish intervention thresholds before clinically apparent neuropathy develops.

Third, our analysis of statistical outliers suggests confounding factors. Two patients with unexpectedly preserved nerve function showed high physical activity levels. These observations align with emerging evidence that physical activity promotes neurological health in individuals with T1D.(53) Future studies should systematically incorporate these factors to develop more comprehensive predictive models.

### 4.5 Conclusion

Despite our study’s null findings, several considerations for clinical practice emerge. First, our observation of low aNCV in this relatively young cohort underscores the importance of early neurological assessment in pediatric T1D.

The suboptimal glycemic control observed despite CGM access highlights persistent challenges in pediatric diabetes management. Our data do not support implementing complex glucose variability metrics over the established glucose SD in clinical practice. Given the lack of meaningful predictive improvement, absence of established reference ranges, and additional training burden for healthcare providers, implementation of advanced variability metrics in routine care appears unjustified based on our findings.

Clinicians should continue focusing on well-established metrics like time-in-range, HbA1c, and standard deviation when interpreting CGM data, as their relationship with complications remains better documented. Regular neurophysiological assessment should be considered for pediatric patients, even those with relatively short diabetes duration, given the evidence of early nerve function changes.

## Data Availability

Some data produced in the present study may be available upon reasonable request to the authors, given that all legal, ethical, and other requirements are met.

