## Appendix for "Assessing Glucose Variability Metrics: Nerve Conduction Velocity in Children and Adolescents with Type 1 Diabetes"

### 7 Appendix

#### 7.1 Detailed Information for selected GV Metrics

##### 7.1.1 Continuous Overall Net Glycemic Action 24h (CONGA24)

Continuous Overall Net Glycemic Action (CONGA) was first introduced by McDonnell et al. in 2005 as a measure for "intraday glycemic variation".(18) CONGA is defined as the standard deviation of the difference between two glucose concentrations n-hours apart. The appropriate n parameter can be chosen by the researcher/clinician depending on the study/clinical objective. Common ones include 1h for short-term fluctuations, 2h for snacking or fast-acting insulin, 4h for larger meals or slow-acting insulin, and 12h for basal rates.(54) By convention, the selected time difference between two glucose concentrations is added to the end of the name CONGAn.

$$CONGAn = \sqrt{\frac{\sum_{t=t_1}^{t_{k^*}} (D_t - \overline{D})^2}{k^* - 1}}$$
$$with\ D_t = GR_t - GR_{t-m}$$
$$and\ \overline{D} = \frac{\sum_{t=t_1}^{t_{k^*}} D_t}{k^*}$$
$$and\ k^* = n\ x\ 60min$$

CONGA has also been leveraged extensively in numerous studies ranging from the validation of synthetic CGM data generation (32) to investigating the relationship between physical activity and GV (33,34) or assessing the impact of various treatment interventions like the use of CGMs (35) or changes in insulin (36). A study by Christensen et al. found a statistically significant association between CONGA and distal symmetric polyneuropathy in children with T1D(17).

##### 7.1.2 Average Daily Risk Range (ADRR)

The Average Daily Risk Range (ADRR) is a glucose variability measure designed by Kovatchev et al. to quantify the risk of hypo- and hyperglycemia. Conceptually, it builds on top of the Low Blood Glucose Index (LBGI) and the High Blood Glucose Index (HBGI) and is defined as the average daily sum of LBGI and HBGI.(30) Unlike many other glucose fluctuation metrics, the ADRR first transforms the data logarithmically to overcome the skewness, typical to CGM data (28). The calculation of the risk scores was initially developed for self-monitoring blood glucose (SMBG) (28) and later adapted for CGM data(29).

$$ADRR = \frac{1}{M}\sum_{i=1}^{M}[LR^i + HR^i]$$
$$with\ LR^i = max[rl(x_1^i), \dots, rl(x_n^i)]\ and\ HR^i = max[rh(x_1^i), \dots, rh(x_n^i)]\ for\ day\ i;\ i = 1,2, \dots, M$$
